# Psychometric evaluation and sex measurement invariance of an ECIPQ-based measure of cybervictimisation and cyberperpetration among Kenyan health-professions students: A multicampus cross-sectional study

**DOI:** 10.64898/2026.11.17.26368285

**Authors:** Anthony Kipkoech Koross, Morris Senghor Shisanya, Ezekiel Kandagor, Deborah Kosgei, Valentine Jepkosgei

## Abstract

Cyberbullying research is constrained by inconsistent measurement, and the instruments most widely used were developed and validated in European and Asian school samples rather than in African higher-education settings. This study evaluated the factor structure, reliability, construct validity and sex measurement invariance of a 22-item measure of cybervictimisation and cyberperpetration based on the European Cyberbullying Intervention Project Questionnaire among health-professions students in Kenya. In a multicampus cross-sectional survey conducted between September and November 2025, 406 students (215 women, 191 men) enrolled at Kenya Medical Training College campuses completed 11 victimisation and 11 perpetration items together with measures of depression, anxiety and stress symptoms, problematic social-media use, self-esteem and well-being. Items were modelled as ordered categorical indicators by confirmatory factor analysis with the WLSMV estimator. A one-factor model was compared with a correlated two-factor model; reliability was estimated using ordinal alpha, McDonald’s omega and composite reliability; convergent validity used Spearman correlations with bootstrap intervals and Holm adjustment; and invariance across sex was tested sequentially. Every item showed a pronounced floor effect. The correlated two-factor model fitted markedly better than the one-factor model (scaled difference χ²(1) = 48.06, p <.001) and, after three content-matched residual covariances were released, fitted the data well (CFI =.984, TLI =.982, RMSEA =.049, 90% CI.042 to.056, SRMR =.044). Standardised loadings ranged from.627 to.928, and the latent correlation between dimensions was.849 (95% CI.803 to.895). Omega was.951 for cybervictimisation and.975 for cyberperpetration. Both dimensions correlated positively with distress and problematic social-media use and negatively with self-esteem, but showed negligible correlations with well-being. Configural, threshold and loading invariance across sex were supported. The findings support scoring the instrument as two reliable, sex-comparable but strongly overlapping dimensions.

## Introduction

Digitally mediated communication now underpins the academic, professional and social lives of young adults, and with it has come sustained concern about aggression carried out through digital media. Cyberbullying is generally understood as intentional and repeated aggressive behaviour enacted through electronic means against a target who finds it difficult to defend themselves [1], and it is conventionally separated into cybervictimisation, the experience of being targeted, and cyberperpetration, the enactment of such behaviour towards others [2]. The two roles are neither mutually exclusive nor independent: a substantial minority of young people report both, and the resulting bully–victim group is consistently associated with the poorest psychosocial profile [3]. Because cyberbullying is associated with depressive and anxiety symptoms, lowered self-esteem and impaired academic functioning [4,5], the credibility of the evidence base rests on whether the instruments used to quantify it measure what they claim to measure in the populations to which they are applied.

That condition is not reliably met. Systematic reviews of cyberbullying assessment repeatedly document substantial heterogeneity in how the construct is operationalised: instruments differ in whether a definition is presented to respondents, in the reference period (from the past month to the whole of life), in whether the response format is binary or frequency-based, in the threshold used to classify involvement, and in whether repetition and power imbalance are represented at all [6,7]. These differences are not merely stylistic. They generate prevalence estimates that vary several-fold across studies, they make cross-study and cross-country comparison hazardous, and they leave open the possibility that apparent group differences reflect differential item functioning rather than genuine differences in the underlying behaviour [7]. Progress therefore depends less on the proliferation of new scales than on rigorous evaluation of existing ones in new populations.

The European Cyberbullying Intervention Project Questionnaire (ECIPQ) was developed to address precisely this problem. It comprises 22 items, 11 addressing victimisation and 11 addressing perpetration, arranged as matched pairs so that each behaviour is asked about from both perspectives. Items are answered on a five-point ordered frequency scale ranging from never to more than once a week within a defined reference period. The original validation, in 5,679 secondary school students from six European countries, supported a two-dimensional conceptualisation and reported comparable factor structures across the national subsamples [8]. The paired structure is a deliberate design feature that supports direct comparison of the two roles, but it also creates the conditions for shared method variance between matched items, an issue that becomes visible only when residual structure is examined explicitly.

Subsequent psychometric work has broadly supported a correlated two-factor structure while diverging on important details. Validations in Colombian and Chinese adolescents reproduced the two dimensions, although the Chinese adaptation retained only 14 of the 22 items after cultural review [9,10]. A large Spanish study likewise supported two factors but reported markedly unequal reliability for the two subscales and established only configural invariance across gender [11], and a subsequent brief form combined cyberbullying items with traditional bullying items [12]. Reported factor correlations vary considerably, and studies differ in whether items were modelled as ordered categorical indicators using polychoric correlations or treated as continuous. This distinction matters: with strongly skewed, low-base-rate items, treating ordered categories as continuous attenuates loadings and distorts fit statistics [13,14], so evidence from the two modelling traditions is not directly comparable.

Evidence from African higher-education settings remains limited. Most ECIPQ validation work has been conducted with school-aged adolescents in Europe, Latin America and East Asia. Kenyan research on cyberbullying has established that the phenomenon is common among university students and has described the forms it takes [15,16], but published studies have generally relied on locally constructed item sets or on instruments used without formal psychometric evaluation in the sampled population. Health-professions students are a particularly relevant group: they are emerging adults in demanding training environments, they use digital platforms intensively for both study and social life, and their psychological well-being has direct implications for the future health workforce [17]. We are not aware of published evidence on the measurement properties of the ECIPQ in Kenyan health-professions students, although the absence of such evidence cannot be established with certainty from the published literature.

Sex is among the most frequently examined correlates of cyberbullying involvement, and meta-analytic evidence suggests small but systematic differences that vary with age [18]. Any such comparison presupposes that the instrument functions equivalently in women and men. Measurement invariance formalises this requirement: configural invariance establishes that the same latent constructs are represented by the same items in both groups, threshold invariance that the points at which respondents move between response categories are comparable, and loading invariance that items relate to the latent variables with the same strength [19–21]. Where these conditions do not hold, observed differences between women and men confound genuine differences in behaviour with differences in how the items are answered. For ordered categorical indicators the sequence and its identification conditions differ from the familiar continuous case, and require the thresholds to be handled explicitly [22,23].

The present study therefore set out to evaluate the factor structure, reliability, construct validity and measurement invariance by sex of the 22-item ECIPQ-based instrument administered to health-professions students at Kenya Medical Training College. The specific objectives were to describe the item-response distributions; to evaluate the factorability of the item set; to compare a one-factor with a correlated two-factor measurement model; to estimate the reliability of each dimension; to evaluate convergent validity against measures of psychological distress, problematic social-media use, self-esteem and well-being; to evaluate discriminant validity between the two dimensions; to test measurement invariance across female and male students; and to determine whether the complete 22-item set could be retained. We expected the items to be strongly skewed towards the lowest response category, the correlated two-factor model to outperform a single-factor specification, both dimensions to reach acceptable reliability, both dimensions to correlate positively with distress and problematic social-media use and negatively with self-esteem and well-being, and at least configural and loading invariance to be supported across sex.

## Materials and methods

### Study design, setting and reporting

This was a psychometric analysis nested within a multicampus cross-sectional survey of students at Kenya Medical Training College (KMTC), a state corporation that is the largest middle-level health training institution in Kenya and delivers pre-service and in-service training in nursing, clinical sciences, public health, diagnostic sciences, pharmaceutical sciences and related fields across campuses distributed nationally. Data were collected between 8 September and 1 November 2025. Participants were drawn from campuses in the eight KMTC sports zones used as the sampling frame (Port Reitz, Manza, Imenti, Nyeri, Kaptumo, Vihiga, Migori and Molo) together with Kabarnet campus; a small number of respondents reported other campuses. Because this manuscript addresses the performance of the instrument rather than the epidemiology of cyberbullying, campus and faculty composition are not analysed or tabulated here. Reporting follows the STROBE statement for cross-sectional studies [24] (S1 Checklist), with additional reference to COSMIN terminology for measurement properties [25,26] and to the SAMPL recommendations for statistical reporting [27].

### Participants and sampling

The target population was students enrolled in health-professions training programmes at the participating KMTC campuses. Students were eligible if they were aged 18 years or older, had completed at least one semester of study, and gave consent. The parent study used multistage cluster sampling in which the eight KMTC sports zones served as clusters, one campus was selected at random from each zone, and the target number of participants was allocated across campuses in proportion to campus enrolment; students within campuses were then approached at random. The sample size for the parent survey was determined using the Yamane formula for a finite population at 95% confidence and 5% precision, giving 377 participants, inflated by 20% to 452 to allow for non-response. A total of 406 eligible students consented and provided complete responses to all 22 instrument items, and this full set was used for every analysis reported here. No separate sample-size calculation was performed for the psychometric analysis, and no retrospective power calculation is reported. The adequacy of a sample for ordinal confirmatory factor analysis depends jointly on the number of indicators, the number of factors, the magnitude of the loadings, the distribution of responses across categories, model complexity, group size and the estimator [28]; these considerations are returned to in the Discussion, particularly in relation to the multigroup models, where the female and male subsamples comprised 215 and 191 participants respectively.

### Data collection

The questionnaire was self-administered electronically using KoBo Toolbox. Participants received a link to the instrument, which opened with a participant information page describing the purpose of the study, the voluntary nature of participation, the right to withdraw at any point without consequence, the confidentiality safeguards in place and the arrangements for referral to the college chaplain, the counselling department or the nearest health facility should any participant become distressed. Consent was recorded electronically by the participant confirming agreement before any study item was displayed; all 406 records in the analytic dataset carry an affirmative consent response. No names, admission numbers or other direct identifiers were collected, and records were identified only by an automatically generated study number. Research assistants were drawn from student support services at the participating campuses and received two days of online training covering use of the electronic data-capture tool, ethical conduct, data management and security, and the counselling and referral pathway. Electronic capture enforced permitted response ranges at the point of entry. The instrument was administered in English, the language of instruction at KMTC.

### Measures

#### Cyberbullying instrument

The instrument comprised 22 items — 11 addressing cybervictimisation and 11 addressing cyberperpetration — based on the European Cyberbullying Intervention Project Questionnaire [8]. The 11 items of each dimension form matched pairs covering the same 11 behavioural content domains from the target and actor perspectives: insult and name-calling, threats, denigration to others, causing fear, password misuse and impersonation, rumour spreading, sharing embarrassing material, deliberate exclusion from online groups, offensive private messages, public ridicule, and identity misuse. Each item was answered on a five-point ordered scale: 0 = never, 1 = once or twice, 2 = once or twice a month, 3 = once a week, 4 = more than once a week. Higher categories therefore indicate more frequent behaviour. Dimension totals range from 0 to 44, with higher scores indicating greater involvement. The full administered wording of every item is reproduced in S6 Table.

The correspondence between the administered items and the published ECIPQ warrants explicit statement. The internal content architecture of the administered instrument — 11 matched victimisation–perpetration pairs spanning the content domains characteristic of the ECIPQ — is fully documented in the study records and is reproduced in S13 Table. However, the source version used, the permissions obtained and any translation, back-translation, expert-panel review, cognitive interviewing or pilot-testing procedures are not documented in the materials available for this analysis. We therefore make no claim that the administered items are semantically equivalent to the published ECIPQ, and we do not describe the instrument as a formally translated or cross-culturally adapted version [29]. Throughout this paper the measure is described as ECIPQ-based. The 22 items were treated as ordered categorical indicators in all factor analyses, and the two dimension totals were used in the external-validity analyses.

#### External construct measures

Depression, anxiety and stress symptoms were assessed with the 21-item Depression Anxiety Stress Scales (DASS-21), which comprises three seven-item subscales rated from 0 to 3 over the preceding week and scaled to a 0–42 range for each domain, with higher scores indicating greater symptom severity [30,31]. DASS-21 scores are treated here as dimensional measures of symptom severity and psychological distress; they are not diagnostic categories and no diagnostic interpretation is made. Problematic social-media use was assessed with the six-item Bergen Social Media Addiction Scale, each item rated from 1 (very rarely) to 5 (very often), giving a total from 6 to 30, with higher scores indicating more problematic use [32,33]. Consistent with the dimensional interpretation adopted here, elevated scores are described as problematic social-media use or elevated risk rather than as a diagnosis of addiction. Self-esteem was assessed with the 10-item Rosenberg Self-Esteem Scale, with the five negatively worded items reverse-scored and a total ranging from 0 to 30, higher scores indicating higher self-esteem [34]. Well-being was assessed with the five-item World Health Organization Well-Being Index, rated from 0 to 5 over the preceding two weeks, summed and multiplied by four to give a percentage score from 0 to 100, higher scores indicating better well-being [35]. All four external measures were analysed as continuous total scores.

#### Data management

Each record carried a unique study identifier. Before analysis the prepared datasets were verified against prespecified expectations: record count, uniqueness of identifiers, absence of duplicates, sex distribution, absence of missing item responses, item values within the permitted 0–4 range, correct assignment of 11 items to each dimension, and exact correspondence between the five-category dataset and a four-category dataset in which the two highest categories were combined. The results of these checks are reported in S1 Table. Involvement-role classifications were held in separate datasets and played no part in any analysis reported here, so that the measurement analysis is independent of any classification rule. Responses were complete for all 22 items and all four external measures in all 406 participants, so no missing-data procedure was required.

#### Item-level analysis

Each item was summarised by the valid number of responses, the frequency and percentage in each response category, the median and interquartile range, the mean and standard deviation for descriptive comparability only, skewness and the corrected item–total correlation with the remaining items of the same dimension. A floor effect was defined as the percentage of participants selecting the lowest category and a ceiling effect as the percentage selecting the highest category, with values above 15% conventionally regarded as notable [36]. A response category was defined as sparse when fewer than five participants selected it, either overall or within a sex group. Categories were recorded as observed overall and separately in women and men. The five original ordered categories were retained for all principal analyses; the four-category coding was used only for the prespecified sensitivity analyses, including the operational check on measurement invariance.

#### Factorability and factor retention

The polychoric correlation matrix of the 22 items was estimated by the two-step procedure of Olsson, with thresholds estimated from the marginal distributions and each correlation by bivariate maximum likelihood [37]. Factorability was assessed using the overall and item-level Kaiser–Meyer–Olkin measures of sampling adequacy [38] and Bartlett’s test of sphericity [39], both computed on the polychoric matrix. The number of dimensions suggested by the data was examined by parallel analysis [40] adapted to ordinal data [41]: each item column was independently permuted, which preserves the observed marginal distributions while removing between-item association, and the polychoric matrix and its eigenvalues were recomputed for each of 1,000 permuted datasets (random seed 20260801). A factor was retained where its observed eigenvalue exceeded the 95th percentile of the corresponding permutation distribution. Parallel analysis was treated as supplementary evidence and did not replace the confirmatory model comparison.

#### Confirmatory factor analysis

Confirmatory models treated all 22 items as ordered categorical indicators and were estimated by diagonally weighted least squares with robust standard errors and a mean-and variance-adjusted (scaled-shifted) test statistic (WLSMV) [42,43]. The delta parameterisation was used with latent variances fixed at one, so that standardised loadings and the factor correlation are reported on the latent-response metric and the residual variance of each latent response variable equals one minus its squared standardised loading.

Analyses were conducted in R version 4.3.3 (R Foundation for Statistical Computing, Vienna, Austria) using lavaan 0.6-17 for polychoric correlations, confirmatory factor analysis and multigroup models, semTools 0.5-6 for measurement-invariance syntax and the heterotrait–monotrait ratio, and psych 2.4.1 for the Kaiser–Meyer–Olkin statistic and Bartlett’s test. The complete analysis code is provided in S1 File, and the reproducibility record, including random seeds and every analytical decision, is provided in S14 Table.

Three models were specified. Model 1 loaded all 22 items on a single general cyberbullying involvement factor. Model 2, the prespecified structure, loaded the 11 victimisation items on a cybervictimisation factor and the 11 perpetration items on a cyberperpetration factor, allowed the two factors to correlate, and fixed all cross-loadings and all residual covariances to zero. Model 3 was a theory-adjusted version of Model 2 in which a small number of residual covariances were released. Model 3 was fitted after estimation of Model 2 and is reported as a post-estimation, theory-supported modification rather than as a prespecified model. A residual covariance was released only where the modification index was among the largest observed, the two items shared documented behavioural content, and a maximum of three modifications in total was respected. For each released parameter the item pair, the modification index, the estimated residual correlation and the content justification are reported in S11 Table.

#### Model fit and item retention

Model fit was evaluated using the scaled chi-square statistic, the comparative fit index (CFI), the Tucker–Lewis index (TLI), the root mean square error of approximation (RMSEA) with its 90% confidence interval, and the standardised root mean square residual (SRMR), together with inspection of standardised loadings, residual variances, the factor correlation and any improper estimates. Conventional benchmarks (CFI and TLI at or above.95 strong and.90 minimally acceptable; RMSEA at or below.06 strong and.08 minimally acceptable; SRMR at or below.06 strong and.08 minimally acceptable) were treated as interpretive guidelines rather than as automatic pass–fail rules [44,45], recognising that these benchmarks were derived for continuous data and that indices from categorical estimators tend to be optimistic. Nested models were compared using the scaled chi-square difference test appropriate to the estimator; unadjusted chi-square values were not subtracted directly [46]. Item retention was judged jointly on loading magnitude and its confidence interval, residual variance, the corrected item–total correlation, conceptual relevance and content coverage, content redundancy, local dependence, stability across sex, and the sensitivity analyses. Items were not deleted solely to improve global fit, and removal was considered only where several diagnostics agreed.

### Reliability

Reliability was estimated separately for each dimension. Ordinal Cronbach’s alpha was computed as standardised alpha from the polychoric correlation matrix of the items of that dimension [47–49]. McDonald’s omega and composite reliability were computed from the standardised loadings of the accepted model, with omega additionally incorporating the released within-dimension residual covariance [50]. Average variance extracted was computed as the mean squared standardised loading. Corrected item–total correlations and alpha with each item removed were also computed. Confidence intervals for omega, composite reliability and average variance extracted were delta-method intervals obtained by defining these quantities as functions of the model parameters, and the interval for ordinal alpha was a percentile bootstrap interval from 1,000 case-resampling replications (seed 20260801). Omega was treated as the principal reliability coefficient, and a combined 22-item alpha is deliberately not reported as a headline value because the instrument is two-dimensional and a single coefficient would misrepresent that structure [51,52].

### Convergent and discriminant validity

Convergent validity was evaluated against six external measures with prespecified directions: positive associations of both ECIPQ dimensions with depression, anxiety and stress symptoms and with problematic social-media use, and negative associations with self-esteem and with well-being. Associations were quantified using Spearman’s rank correlation between each dimension total and each external score, with percentile bootstrap 95% confidence intervals from 2,000 case-resampling replications using a fixed random seed of 20260801; all 2,000 replications were valid. Tests were two-sided and p-values were adjusted using the Holm procedure across the six external measures separately within each dimension [53,54]. All correlations used the complete sample of 406 participants, since no external measure had missing values.

Discriminant validity between cybervictimisation and cyberperpetration was assessed using five converging approaches rather than any single criterion: the scaled difference test comparing the one-factor and correlated two-factor models; the magnitude of the latent factor correlation; the confidence interval for that correlation and whether it excluded unity; the Fornell–Larcker comparison of the squared latent correlation with each dimension’s average variance extracted [55]; the heterotrait–monotrait ratio computed from the polychoric matrix [56]; and the observed-score correlation between the two dimension totals. No single threshold was treated as decisive.

### Measurement invariance by sex

Invariance across female and male students was examined in a sequence appropriate to ordered categorical indicators [22,23]. Separate single-group models were first fitted in women and in men to confirm that the accepted structure was interpretable within each group. Multigroup models were then identified following Wu and Estabrook, which is designed for ordered categorical indicators and avoids the untestable constraints implied by conventional identification: a configural model with the same pattern of loadings in both groups and with loadings, thresholds and factor correlations freely estimated; a threshold-invariance model in which all item thresholds were constrained equal across sex; and a model constraining both thresholds and loadings. Residual (strict) invariance was not tested because it is not required for comparing latent means. Each added level of constraint was evaluated with the scaled chi-square difference test and with working criteria of a change in CFI no worse than −.010 and an increase in RMSEA of no more than.015 [57,58]; a significant difference test alone was not treated as sufficient grounds for rejecting invariance [59]. Partial invariance would have been considered had any parameter been non-invariant, retaining at least two invariant indicators per factor. Latent means were compared only after threshold and loading invariance had been established, with the female group as reference (latent mean fixed at zero, latent variance fixed at one). Because three item categories were unobserved among women under the five-category coding, and multigroup estimation requires every category to be observed in each group, the invariance models used the prespecified four-category coding, in which the two highest categories were combined and no category was empty in either sex.

### Sensitivity analyses

Eight sensitivity analyses were specified. Prespecified analyses compared the five-category with the four-category coding, the prespecified with the theory-adjusted model specification, ordinal with continuous (Pearson) treatment of the items using robust maximum likelihood, and the three reliability estimators. Conditional analyses covered exclusion of exceptional response patterns. Exploratory analyses covered exclusion of participants who scored zero on every item and estimation of a supplementary bifactor model. Two prespecified analyses were not undertaken and the reasons are stated: an exploratory refined item set was not estimated because the prespecified condition for item deletion was never met, and latent external-validity correlations were not modelled because the external measures were available only as total scores, so that a latent model would have required single-indicator constructs with assumed reliabilities. Results are summarised in S10 Table.

### Ethical considerations

The study was conducted in accordance with the Declaration of Helsinki [60]. The protocol was approved by the Institutional Scientific and Ethics Review Committee of Masinde Muliro University of Science and Technology (MMUST-ISERC; approval number MMUST/ISERC/091/2025, 2 July 2025). A research licence was issued by the National Commission for Science, Technology and Innovation (NACOSTI; licence NACOSTI/P/25/4176509, 29 July 2025), and the Kenya Medical Training College granted institutional permission for data collection at its campuses after review by the College’s Research and Ethics Review Committee (reference KMTC/ADM/74/VOL.VI(614), 11 August 2025). All participants were adults aged 18 years or older. Informed consent was obtained electronically: participants read the participant information and confirmed their agreement within the electronic questionnaire before any study item was presented; written signed consent was not obtained because the questionnaire was self-administered remotely, and the electronic consent procedure was approved by the ethics committee. Participation was voluntary and participants could decline or withdraw at any point without consequence. No direct identifiers were collected, data were held on password-protected systems accessible only to the research team, and participants who reported distress were offered referral to campus counselling services, the campus chaplain or the nearest health facility.

### Use of artificial intelligence tools

Anthropic’s Claude AI was used in 2026 to assist with counterchecking the R analysis code and language editing of manuscript text. The authors take full responsibility for the content of the article.

## Results

### Participants and data completeness

All 406 eligible, consenting students provided complete responses to all 22 instrument items and to all four external measures, so every analysis used the full sample and no missing-data procedure was required. Verification of the analysis inputs matched every prespecified expectation, including unique identifiers with no duplicates, item values within the permitted 0–4 range, correct assignment of 11 items to each dimension and exact correspondence between the five-category and four-category datasets (S1 Table). Participants had a mean age of 21.51 years (SD 2.95, median 21.0, interquartile range 20.0–23.0, range 18–41); 215 (53.0%) were women and 191 (47.0%) were men; and 126 (31.0%) were in year 1, 196 (48.3%) in year 2 and 84 (20.7%) in year 3 or above. Characteristics of the psychometric sample and the item-response distributions are given in Table 1. A total of 128 participants (31.5%) responded zero to all 22 items; these are valid uninvolved respondents, were retained throughout, and were examined separately in a sensitivity analysis.

**Table 1.**
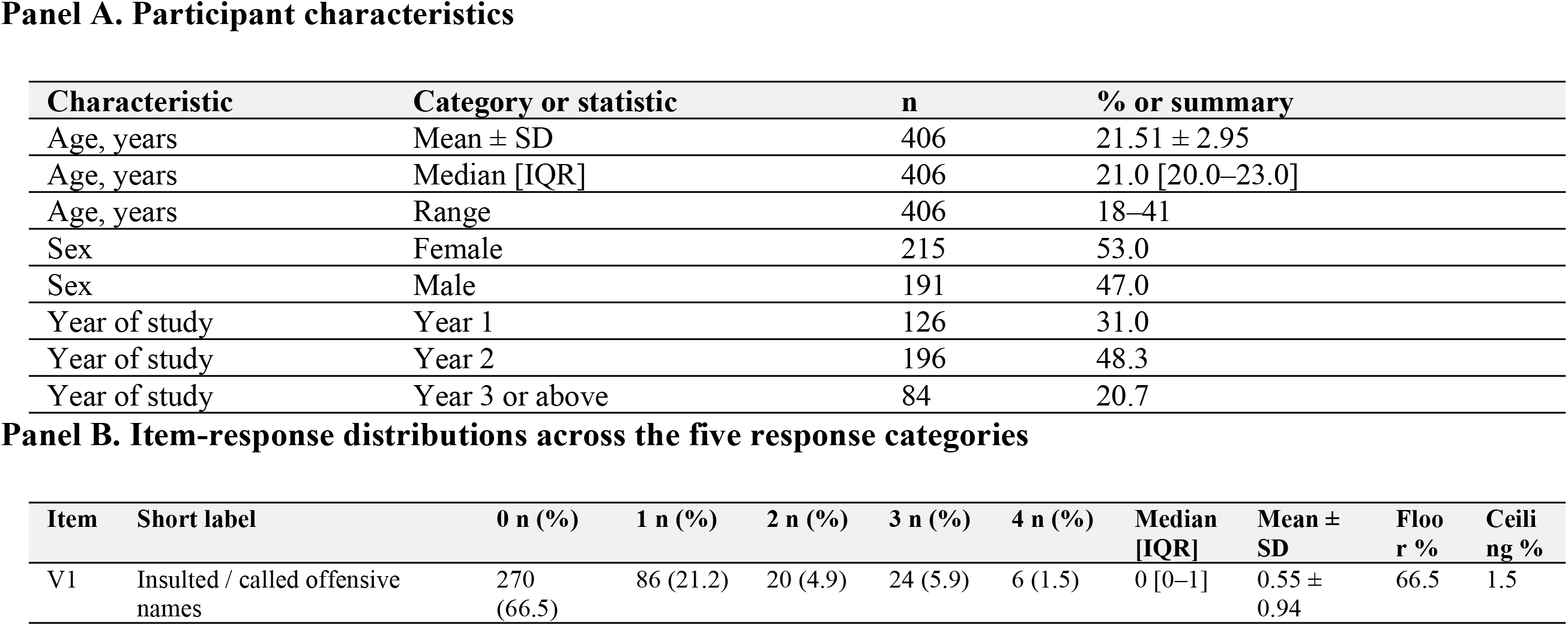
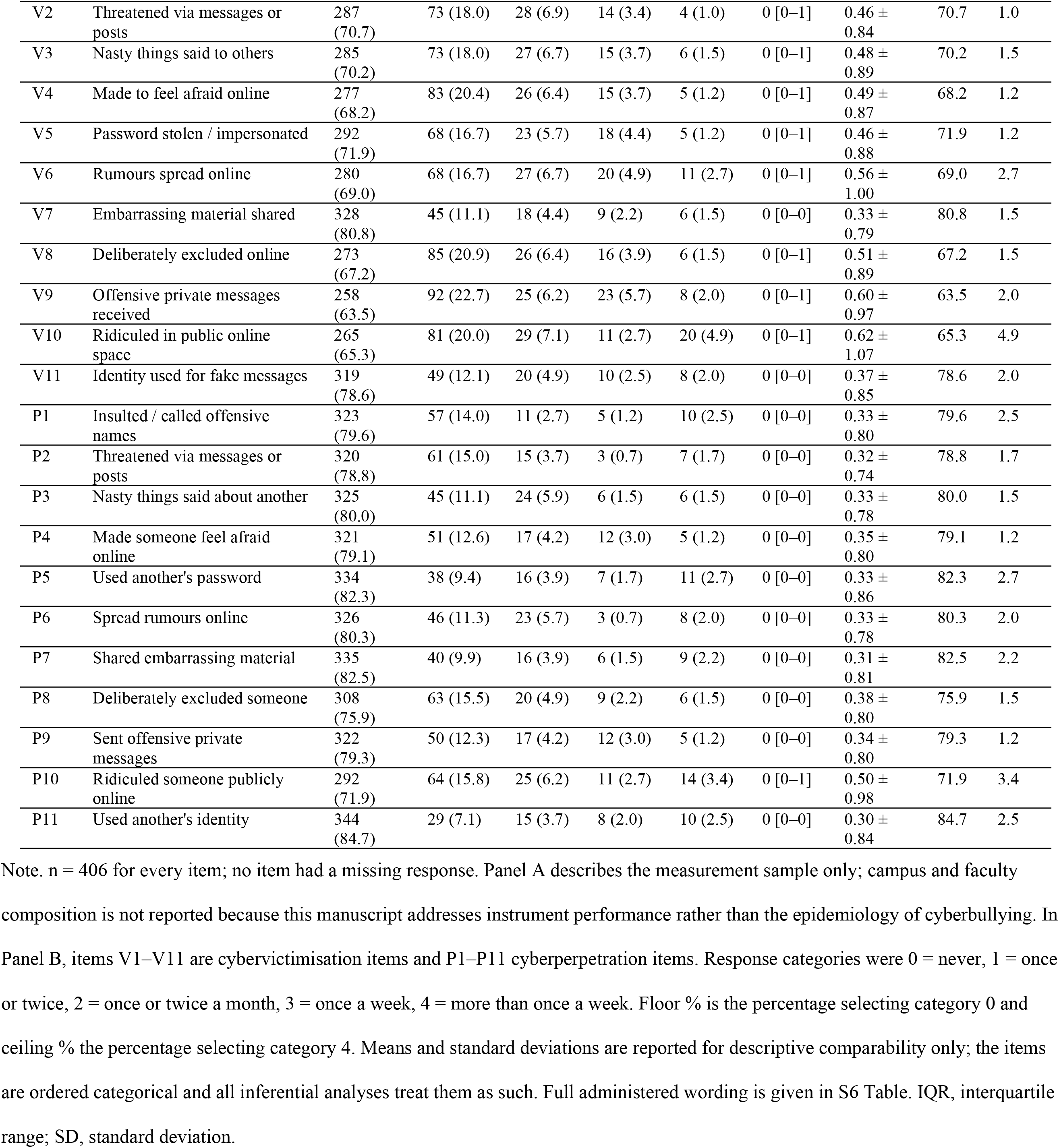
Participant characteristics and ECIPQ item-response distributions.

| Panel A. Participant characteristics |  |  |  |  |  |  |  |  |  |
| Characteristic | Category or statistic | n | % or summary |  |  |  |  |  |  |
|---|---|---|---|---|---|---|---|---|---|
| Age, years | Mean ± SD | 406 | 21.51 ± 2.95 |  |  |  |  |  |  |
| Age, years | Median [IQR] | 406 | 21.0 [20.0–23.0] |  |  |  |  |  |  |
| Age, years | Range | 406 | 18–41 |  |  |  |  |  |  |
| Sex | Female | 215 | 53.0 |  |  |  |  |  |  |
| Sex | Male | 191 | 47.0 |  |  |  |  |  |  |
| Year of study | Year 1 | 126 | 31.0 |  |  |  |  |  |  |
| Year of study | Year 2 | 196 | 48.3 |  |  |  |  |  |  |
| Year of study | Year 3 or above | 84 | 20.7 |  |  |  |  |  |  |
| **Panel B. Item-response distributions across the five response categories** |  |  |  |  |  |  |  |  |  |
| **Item** | **Short label** | **0 n (%)** | **1 n (%)** | **2 n (%)** | **3 n (%)** | **4 n (%)** | **Median [IQR]** | **Mean ± SD** | **Floor %** | **Ceiling %** |
| V1 | Insulted / called offensive names | 270 (66.5) | 86 (21.2) | 20 (4.9) | 24 (5.9) | 6 (1.5) | 0 [0–1] | 0.55 ± 0.94 | 66.5 | 1.5 |
| V2 | Threatened via messages or posts | 287 (70.7) | 73 (18.0) | 28 (6.9) | 14 (3.4) | 4 (1.0) | 0 [0–1] | 0.46 ± 0.84 | 70.7 | 1.0 |
| V3 | Nasty things said to others | 285 (70.2) | 73 (18.0) | 27 (6.7) | 15 (3.7) | 6 (1.5) | 0 [0–1] | 0.48 ± 0.89 | 70.2 | 1.5 |
| V4 | Made to feel afraid online | 277 (68.2) | 83 (20.4) | 26 (6.4) | 15 (3.7) | 5 (1.2) | 0 [0–1] | 0.49 ± 0.87 | 68.2 | 1.2 |
| V5 | Password stolen / impersonated | 292 (71.9) | 68 (16.7) | 23 (5.7) | 18 (4.4) | 5 (1.2) | 0 [0–1] | 0.46 ± 0.88 | 71.9 | 1.2 |
| V6 | Rumours spread online | 280 (69.0) | 68 (16.7) | 27 (6.7) | 20 (4.9) | 11 (2.7) | 0 [0–1] | 0.56 ± 1.00 | 69.0 | 2.7 |
| V7 | Embarrassing material shared | 328 (80.8) | 45 (11.1) | 18 (4.4) | 9 (2.2) | 6 (1.5) | 0 [0–0] | 0.33 ± 0.79 | 80.8 | 1.5 |
| V8 | Deliberately excluded online | 273 (67.2) | 85 (20.9) | 26 (6.4) | 16 (3.9) | 6 (1.5) | 0 [0–1] | 0.51 ± 0.89 | 67.2 | 1.5 |
| V9 | Offensive private messages received | 258 (63.5) | 92 (22.7) | 25 (6.2) | 23 (5.7) | 8 (2.0) | 0 [0–1] | 0.60 ± 0.97 | 63.5 | 2.0 |
| V10 | Ridiculed in public online space | 265 (65.3) | 81 (20.0) | 29 (7.1) | 11 (2.7) | 20 (4.9) | 0 [0–1] | 0.62 ± 1.07 | 65.3 | 4.9 |
| V11 | Identity used for fake messages | 319 (78.6) | 49 (12.1) | 20 (4.9) | 10 (2.5) | 8 (2.0) | 0 [0–0] | 0.37 ± 0.85 | 78.6 | 2.0 |
| P1 | Insulted / called offensive names | 323 (79.6) | 57 (14.0) | 11 (2.7) | 5 (1.2) | 10 (2.5) | 0 [0–0] | 0.33 ± 0.80 | 79.6 | 2.5 |
| P2 | Threatened via messages or posts | 320 (78.8) | 61 (15.0) | 15 (3.7) | 3 (0.7) | 7 (1.7) | 0 [0–0] | 0.32 ± 0.74 | 78.8 | 1.7 |
| P3 | Nasty things said about another | 325 (80.0) | 45 (11.1) | 24 (5.9) | 6 (1.5) | 6 (1.5) | 0 [0–0] | 0.33 ± 0.78 | 80.0 | 1.5 |
| P4 | Made someone feel afraid online | 321 (79.1) | 51 (12.6) | 17 (4.2) | 12 (3.0) | 5 (1.2) | 0 [0–0] | 0.35 ± 0.80 | 79.1 | 1.2 |
| P5 | Used another's password | 334 (82.3) | 38 (9.4) | 16 (3.9) | 7 (1.7) | 11 (2.7) | 0 [0–0] | 0.33 ± 0.86 | 82.3 | 2.7 |
| P6 | Spread rumours online | 326 (80.3) | 46 (11.3) | 23 (5.7) | 3 (0.7) | 8 (2.0) | 0 [0–0] | 0.33 ± 0.78 | 80.3 | 2.0 |
| P7 | Shared embarrassing material | 335 (82.5) | 40 (9.9) | 16 (3.9) | 6 (1.5) | 9 (2.2) | 0 [0–0] | 0.31 ± 0.81 | 82.5 | 2.2 |
| P8 | Deliberately excluded someone | 308 (75.9) | 63 (15.5) | 20 (4.9) | 9 (2.2) | 6 (1.5) | 0 [0–0] | 0.38 ± 0.80 | 75.9 | 1.5 |
| P9 | Sent offensive private messages | 322 (79.3) | 50 (12.3) | 17 (4.2) | 12 (3.0) | 5 (1.2) | 0 [0–0] | 0.34 ± 0.80 | 79.3 | 1.2 |
| P10 | Ridiculed someone publicly online | 292 (71.9) | 64 (15.8) | 25 (6.2) | 11 (2.7) | 14 (3.4) | 0 [0–1] | 0.50 ± 0.98 | 71.9 | 3.4 |
| P11 | Used another's identity | 344 (84.7) | 29 (7.1) | 15 (3.7) | 8 (2.0) | 10 (2.5) | 0 [0–0] | 0.30 ± 0.84 | 84.7 | 2.5 |

Note. n = 406 for every item; no item had a missing response. Panel A describes the measurement sample only; campus and faculty composition is not reported because this manuscript addresses instrument performance rather than the epidemiology of cyberbullying. In Panel B, items V1–V11 are cybervictimisation items and P1–P11 cyberperpetration items. Response categories were 0 = never, 1 = once or twice, 2 = once or twice a month, 3 = once a week, 4 = more than once a week. Floor % is the percentage selecting category 0 and ceiling % the percentage selecting category 4. Means and standard deviations are reported for descriptive comparability only; the items are ordered categorical and all inferential analyses treat them as such. Full administered wording is given in S6 Table. IQR, interquartile range; SD, standard deviation.

### Item-response distributions

Every item was strongly positively skewed with a median of 0. The proportion of students reporting that a behaviour had never occurred ranged from 63.5% (offensive private messages received) to 84.7% (used another’s identity), and skewness ranged from 1.77 to 3.11. Floor effects were therefore pronounced throughout and well above the conventional 15% threshold, whereas ceiling effects were uniformly small, ranging from 1.0% to 4.9%; no item exceeded 4.9% in the highest frequency category. Victimisation items were endorsed somewhat more often than perpetration items, with floor percentages of 63.5% to 80.8% and 71.9% to 84.7% respectively. Item means ranged from 0.30 to 0.62. All 22 items used the full 0–4 response range in the total sample, but upper categories were thinly populated: three items had a sparse category (fewer than five respondents) in the total sample, and 20 of the 22 items had at least one sparse category within at least one sex group. Among the 215 women, three item categories were completely unobserved; among the 191 men, no category was empty. This asymmetry shaped the invariance strategy but did not prevent estimation. The complete distributions are shown in Fig 1.

**Fig 1.**
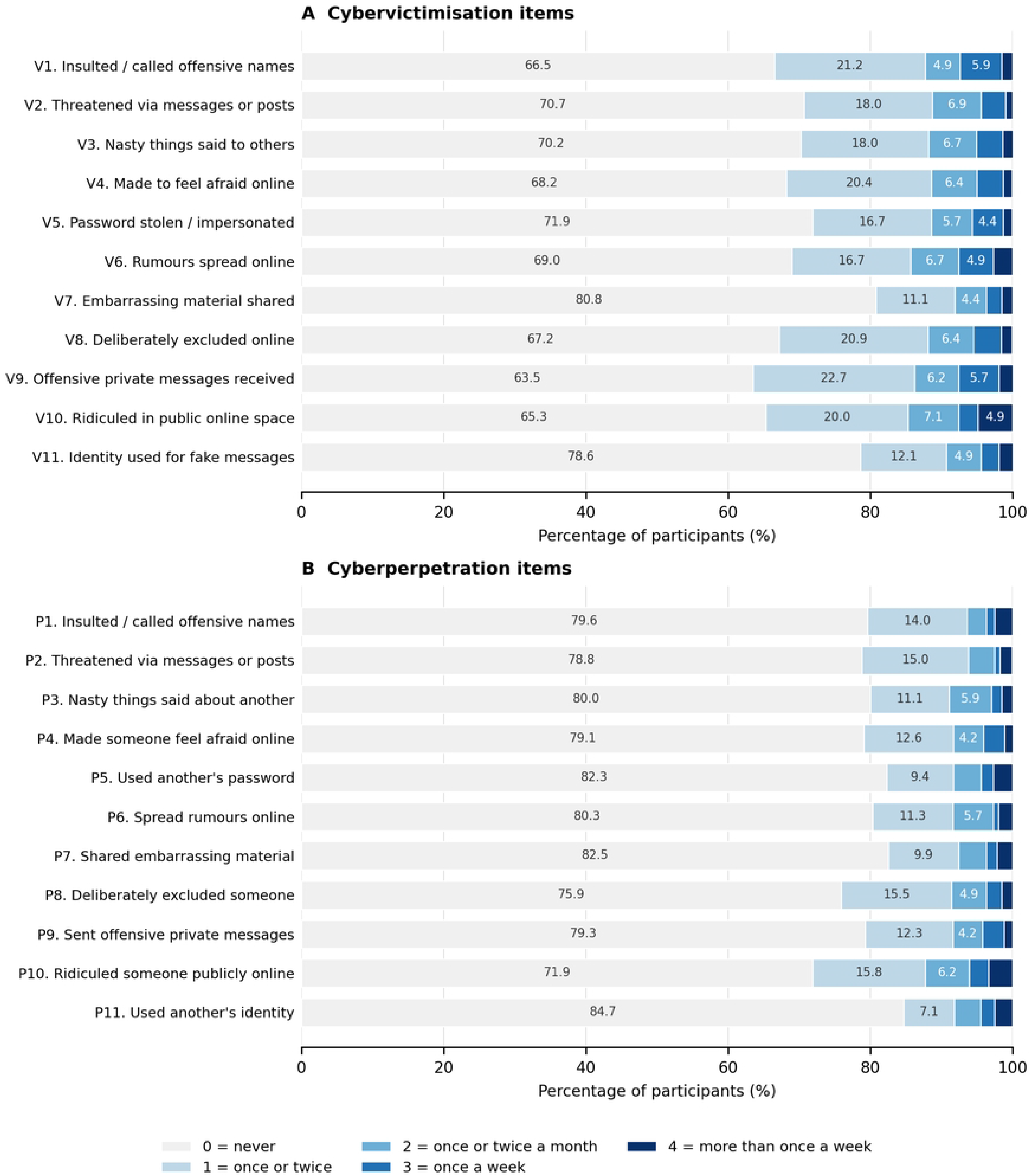
Response-category distributions across the 22 ECIPQ items.

Horizontal 100% stacked bars show the percentage of the 406 participants selecting each response category for every item. Panel A shows the 11 cybervictimisation items and Panel B the 11 cyberperpetration items, each in the administered order with item 1 at the top. Categories are 0 = never, 1 = once or twice, 2 = once or twice a month, 3 = once a week and 4 = more than once a week. Valid n = 406 for every item, with no missing responses. Segment labels are shown where the category accounts for at least 4% of responses. Full item wording is given in S6 Table.

### Factorability and parallel analysis

The polychoric correlation matrix was clearly suitable for factor analysis. The overall Kaiser–Meyer–Olkin measure was.870, item-level values ranged from.645 to.953 and all exceeded the.50 minimum, and Bartlett’s test rejected sphericity, χ²(231) = 11710.9, p <.001 (S2 Table). Polychoric correlations between items ranged from.256 to.890, the matrix was positive definite (smallest eigenvalue.020), and no smoothing or other adjustment was applied (S12 Table). Parallel analysis based on 1,000 permutations retained a single factor: the first observed eigenvalue of 14.751 far exceeded its 95th-percentile permutation reference of 1.903, whereas the second observed eigenvalue of 1.625 did not exceed its reference of 1.730, and the third observed eigenvalue was.726 (S3 Table and S1 Fig). This pattern indicates a strong general involvement dimension underlying both item sets, and is consistent with the high latent correlation estimated in the confirmatory models reported below. Eigenvalue-based criteria are known to under-extract when factors are strongly correlated and cannot test a prespecified structure, so this result is reported alongside, and not in place of, the confirmatory model comparison.

### Confirmatory factor analysis

Comparative fit of the measurement models is given in Table 2. The one-factor model fitted least well (CFI =.954, TLI =.949, RMSEA =.082, 90% CI.076 to.088, SRMR =.074). The prespecified correlated two-factor model fitted better (CFI =.979, TLI =.976, RMSEA =.056, 90% CI.049 to.062, SRMR =.048), and the scaled difference test strongly favoured it (χ²(1) = 48.06, p <.001). Modification indices from the two-factor model identified localised residual dependence, and the three largest all involved items with shared documented content; these residual covariances were released: between the matched pair V10 and P10 (public ridicule), between the matched pair V1 and P1 (insult and name-calling), and between V3 and V6 within the victimisation dimension (denigration to others and rumour spreading, which overlap in content). The resulting theory-adjusted model gave the best fit (CFI =.984, TLI =.982, RMSEA =.049, 90% CI.042 to.056, SRMR =.044) and improved significantly on the prespecified model (χ²(3) = 65.57, p <.001). All models converged, and no model produced an improper estimate, a negative residual variance or a standardised loading above one. The theory-adjusted correlated two-factor model was accepted as the principal model, and four-category models reproduced these results closely. The scaled chi-square remained significant for every model, as is usual at this sample size and model complexity. The recently proposed robust (bias-corrected) versions of CFI and RMSEA for categorical data were less favourable for every model (accepted model CFI =.859, RMSEA =.142); these corrections are known to behave unstably with sparse, highly skewed ordinal items such as these, and they ranked the models in the same order, so they do not alter the model comparison but do caution against treating the conventional indices as evidence of close fit.

**Table 2.** Comparative fit of alternative ECIPQ measurement models.

| Model | Item coding | Scaled χ² | df | p | CFI | TLI | RMSEA | RMSEA 90% CI | SRMR | Factor correlation | Decision |
|---|---|---|---|---|---|---|---|---|---|---|---|
| Model 1: one general involvement factor | Five-category ordered | 776.9 | 209 | < .001 | .954 | .949 | .082 | [.076, .088] | .074 | — (single factor) | Rejected in favour of the two-factor model |
| Model 2: prespecified correlated two-factor model | Five-category ordered | 469.5 | 208 | < .001 | .979 | .976 | .056 | [.049, .062] | .048 | .852 (SE .023; 95% CI [.806, .897]) | Retained as the prespecified structure |
| Model 3: theory-adjusted correlated two-factor model | Five-category ordered | 402.1 | 205 | < .001 | .984 | .982 | .049 | [.042, .056] | .044 | .849 (SE .023; 95% CI [.803, .895]) | Accepted principal model |
| Model 2 (four-category sensitivity) | Four-category ordered (3 and 4 combined) | 429.3 | 208 | < .001 | .985 | .984 | .051 | [.044, .058] | .048 | .850 | Sensitivity: conclusions unchanged |
| Model 3 (four-category sensitivity) | Four-category ordered (3 and 4 combined) | 371.7 | 205 | < .001 | .989 | .987 | .045 | [.037, .052] | .044 | .848 | Sensitivity: conclusions unchanged |
| Supplementary: bifactor model (general plus two specific factors) | Five-category ordered | 429.5 | 187 | < .001 | .980 | .976 | .057 | [.050, .064] | .040 | Orthogonal by specification | Supplementary only; fitted less well than Model 3 |
| Sensitivity: continuous-item model (robust maximum likelihood) | Items treated as continuous | 984.5 | 205 | < .001 | .872 | .856 | .097 | — | .053 | .807 | Illustrative only — not the definitive estimator |
| **Scaled nested model comparisons** |  |  |  |  |  |  |  |  |  |  |  |
| **Comparison** | **Scaled difference χ²** | **Δdf** | **p** | **ΔCFI** | **ΔRMSEA** | **ΔSRMR** | **Preferred model** |  |  |  |  |
| Model 1 (one factor) versus Model 2 (correlated two factors) | 48.06 | 1 | < .001 | .025 | −.026 | −.026 | Model 2 |  |  |  |  |
| Model 2 versus Model 3 (three theory-supported residual covariances) | 65.57 | 3 | < .001 | .005 | −.007 | −.004 | Model 3 |  |  |  |  |
| Model 3 (five-category) versus Model 3 (four-category sensitivity) | Not nested — different item coding | — | — | .005 | −.004 | .000 | Equivalent conclusions |  |  |  |  |

Note. Analysed n = 406 for every model except the continuous-item model, which used the same 406 participants with robust maximum likelihood. Ordinal models were estimated by WLSMV (diagonally weighted least squares with robust standard errors and a mean- and variance-adjusted, scaled-shifted test statistic) in lavaan 0.6-17, using the delta parameterisation; CFI, TLI and RMSEA are the scaled versions. Model 3 releases three theory-supported residual covariances identified from modification indices and confirmed against documented matched content (V10 with P10; V1 with P1; V3 with V6). Nested models were compared with the scaled chi-square difference test; unadjusted chi-square values were not subtracted directly. The five-category and four-category models are fitted to different codings of the same responses and are therefore not nested, so only descriptive differences in fit indices are reported for that comparison. The bifactor and continuous-item rows are supplementary; the continuous-item statistics are robust maximum-likelihood statistics and are not directly comparable with the categorical statistics above them. Interpretive guidelines: CFI and TLI ≥ .95 strong and ≥ .90 minimally acceptable; RMSEA ≤ .06 strong and ≤ .08 minimally acceptable; SRMR ≤ .06 strong and ≤ .08 minimally acceptable.
CFI, comparative fit index; CI, confidence interval; RMSEA, root mean square error of approximation; SE, standard error; SRMR, standardised root mean square residual; TLI, Tucker–Lewis index.

### Factor loadings and item performance

In the accepted model all 22 items loaded substantially and precisely on their intended factor (Table 3, Panel A; Fig 2). Standardised loadings ranged from.627 to.928, every loading was significant at p <.001 with a minimum z of 14.9, and the lowest confidence limit for any loading was.545. For cybervictimisation, loadings ranged from.627 (insulted or called offensive names) to.895 (embarrassing material shared); for cyberperpetration they ranged from.782 (insulted or called someone offensive names) to.928 (sent offensive private messages). Twenty-one of the 22 loadings exceeded.70, and the one item below.70, which also carried one of the largest floor effects, loaded at.627. Corrected item–total correlations ranged from.512 to.803. Removing any single item never raised ordinal alpha for its dimension above the full-scale value by a material margin (ranges.947 to.954 for victimisation and.970 to.974 for perpetration; S5 Table), and no item met the prespecified multi-criterion threshold for exclusion. All 22 items were therefore retained and no shortened model was estimated. Complete parameter estimates, including all item thresholds, are given in S4 Table.

**Fig 2.**
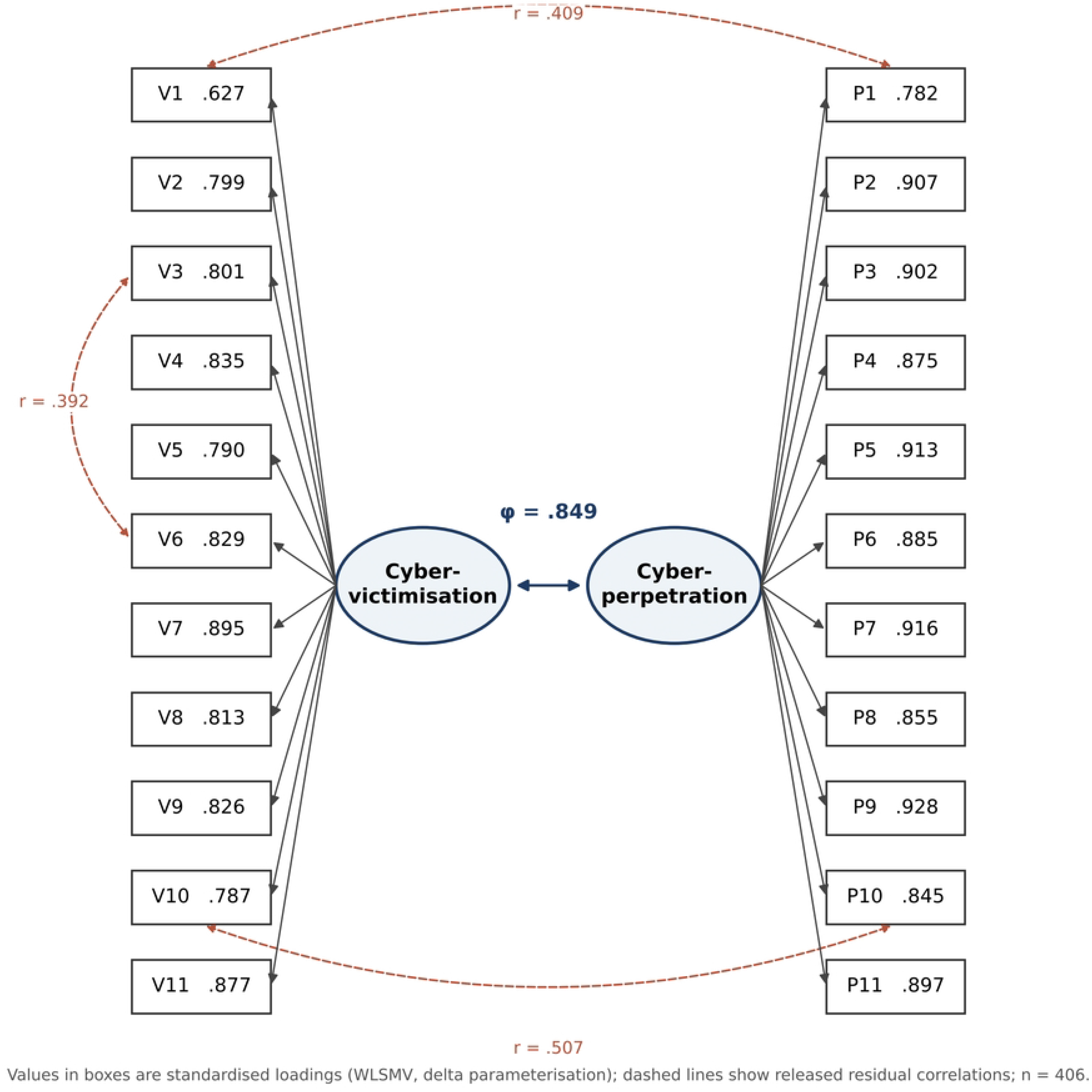
Standardised correlated two-factor model of the ECIPQ-based measure. The accepted theory-adjusted model (n = 406) with two latent factors, 11 indicators each, standardised loadings on the single-headed arrows, and the latent factor correlation on the double-headed arrow between the factors. Dashed double-headed arrows show the three released residual covariances, expressed as residual correlations (V10 with P10, r =.507; V1 with P1, r =.409; V3 with V6, r =.392). Item thresholds are not shown and are reported in S4 Table; full item wording is given in S6 Table. Estimation was by WLSMV with the delta parameterisation.

**Table 3.** Item performance, factor loadings and score reliability in the accepted model.

| **Item** | **Short description** | **Dimension** | **Standardised loading** | **95% CI** | **Residual variance** | **Corrected item–total r** | **Ordinal α if item removed** | **Retention** |
|---|---|---|---|---|---|---|---|---|
| V1 | Insulted / called offensive names | Victimisation | .627 | [.545, .710] | .607 | .512 | .954 | Adequate — retain |
| V2 | Threatened via messages or posts | Victimisation | .799 | [.743, .855] | .362 | .683 | .948 | Strong — retain |
| V3 | Nasty things said to others | Victimisation | .801 | [.744, .858] | .359 | .696 | .948 | Strong — retain |
| V4 | Made to feel afraid online | Victimisation | .835 | [.788, .883] | .302 | .694 | .948 | Strong — retain |
| V5 | Password stolen / impersonated | Victimisation | .790 | [.730, .849] | .377 | .635 | .950 | Strong — retain |
| V6 | Rumours spread online | Victimisation | .829 | [.780, .877] | .313 | .720 | .947 | Strong — retain |
| V7 | Embarrassing material shared | Victimisation | .895 | [.847, .942] | .199 | .685 | .947 | Strong — retain |
| V8 | Deliberately excluded online | Victimisation | .813 | [.763, .864] | .338 | .673 | .949 | Strong — retain |
| V9 | Offensive private messages received | Victimisation | .826 | [.778, .874] | .317 | .728 | .947 | Strong — retain |
| V10 | Ridiculed in public online space | Victimisation | .787 | [.728, .845] | .381 | .628 | .950 | Strong — retain |
| V11 | Identity used for fake messages | Victimisation | .877 | [.826, .927] | .232 | .668 | .948 | Strong — retain |
| P1 | Insulted / called offensive names | Perpetration | .782 | [.717, .847] | .389 | .588 | .974 | Strong — retain |
| P2 | Threatened via messages or posts | Perpetration | .907 | [.872, .941] | .178 | .768 | .970 | Strong — retain |
| P3 | Nasty things said about another | Perpetration | .902 | [.867, .936] | .187 | .772 | .970 | Strong — retain |
| P4 | Made someone feel afraid online | Perpetration | .875 | [.832, .918] | .234 | .705 | .971 | Strong — retain |
| P5 | Used another's password | Perpetration | .913 | [.877, .949] | .167 | .761 | .970 | Strong — retain |
| P6 | Spread rumours online | Perpetration | .885 | [.846, .924] | .217 | .742 | .971 | Strong — retain |
| P7 | Shared embarrassing material | Perpetration | .916 | [.882, .950] | .161 | .779 | .970 | Strong — retain |
| P8 | Deliberately excluded someone | Perpetration | .855 | [.814, .897] | .268 | .719 | .972 | Strong — retain |
| P9 | Sent offensive private messages | Perpetration | .928 | [.900, .956] | .139 | .803 | .970 | Strong — retain |
| P10 | Ridiculed someone publicly online | Perpetration | .845 | [.800, .889] | .286 | .701 | .971 | Strong — retain |
| P11 | Used another's identity | Perpetration | .897 | [.853, .940] | .196 | .735 | .970 | Strong — retain |
| **Panel B. Dimension-level score reliability and average variance extracted** |  |  |  |  |  |  |  |  |
| **Dimension** | **Items** | **Ordinal Cronbach's α [95% CI]** | **McDonald's ω [95% CI]** | **Composite reliability** | **Average variance extracted [95% CI]** | **Observed score range** |  |  |
| Cybervictimisation | 11 | .953 [.942, .963] | .951 [.941, .961] | .954 | .656 [.607, .705] | 0–44 |  |  |
| Cyberperpetration | 11 | .973 [.966, .980] | .975 [.969, .981] | .975 | .780 [.736, .824] | 0–44 |  |  |

Note. Accepted principal model: theory-adjusted correlated two-factor ordinal confirmatory factor analysis estimated by WLSMV with the delta parameterisation, n = 406. Loadings are standardised and the residual variance of each latent response variable equals 1 − λ². Confidence intervals for loadings are Wald intervals from robust standard errors; all loadings were significant at p < .001 (minimum z = 14.9). Corrected item–total correlations are between the item and the sum of the remaining items of the same dimension. Ordinal α if item removed is computed from the polychoric correlation submatrix of the remaining items of that dimension. In Panel B, ordinal α is standardised alpha from the polychoric matrix, with a percentile bootstrap 95% CI from 1,000 replications; ω and composite reliability are computed from the accepted model's standardised loadings, ω additionally incorporating the released within-dimension residual covariance; average variance extracted is the mean squared standardised loading; intervals for ω and average variance extracted are delta-method intervals. ω is the principal reliability coefficient; a combined 22-item α is deliberately not reported as a headline value because the instrument is two-dimensional. Retention guidelines: ≥ .70 strong, .50–.69 adequate, .40–.49 marginal, < .40 potentially weak. No item was removed. CI, confidence interval.

### Reliability

Both dimensions showed high internal consistency (Table 3, Panel B). For cybervictimisation, ordinal alpha was.953 (95% CI.942 to.963), McDonald’s omega.951 (95% CI.941 to.961) and composite reliability.954; for cyberperpetration the corresponding values were.973 (95% CI.966 to.980),.975 (95% CI.969 to.981) and.975. The three estimators agreed to within.003 in both dimensions, so the reliability conclusion does not depend on which coefficient is chosen. Average variance extracted was.656 (95% CI.607 to.705) for cybervictimisation and.780 (95% CI.736 to.824) for cyberperpetration. These coefficients indicate that dimension scores are reliable enough for group-level research use. They should not, however, be read as evidence of unidimensionality or of validity: the values partly reflect the strong general involvement dimension and the concentration of responses at the floor, and the victimisation average variance extracted bears directly on the discriminant-validity evidence considered next.

### Discriminant validity

Evidence on the separability of the two dimensions was mixed but on balance supportive of two related factors (S7 Table). The correlated two-factor model fitted materially better than the one-factor model (scaled difference χ²(1) = 48.06, p <.001; CFI.979 versus.954). The latent factor correlation in the accepted model was.849 (robust SE.023) with a 95% confidence interval of.803 to.895 that excluded 1.00, indicating that the dimensions are strongly but not perfectly related; the magnitude nonetheless approaches the.85 to.90 range at which distinctness is conventionally questioned. The heterotrait–monotrait ratio was.842, below both the.90 and.85 thresholds. The observed-score correlation between the dimension totals was ρ =.655 (bootstrap 95% CI.586 to.717), substantial but far from unity. Against these supportive findings, the Fornell–Larcker criterion was met for cyberperpetration but not for cybervictimisation: the squared latent correlation of.721 exceeded the victimisation average variance extracted of.656 while remaining below the perpetration value of.780. Taken together with the single-factor result from parallel analysis, this indicates that a strong general involvement dimension underlies both subscales, so discriminant validity is classified here as partially supported.

### Convergent validity

Five of the six prespecified convergent-validity hypotheses were supported for both dimensions (Table 4). Both cybervictimisation and cyberperpetration correlated positively and moderately with depression (ρ =.506 and.509), anxiety (.507 and.470) and stress symptoms (.507 and.459), positively with problematic social-media use (.338 and.229), and negatively with self-esteem (−.251 and −.243); every one of these ten associations was in the predicted direction with a confidence interval excluding zero and a Holm-adjusted p <.001. The exception was well-being: correlations with the WHO-5 were negligible and not statistically significant for either dimension (cybervictimisation ρ = −.064, 95% CI −.160 to.032, Holm-adjusted p =.198; cyberperpetration ρ = −.020, 95% CI −.119 to.077, Holm-adjusted p =.695). This null finding is contrary to the prespecified expectation and is reported rather than omitted. Because these are observed-score correlations, attenuated both by measurement error and by the strong floor effects in the instrument, they are conservative estimates of the underlying latent associations.

**Table 4.**
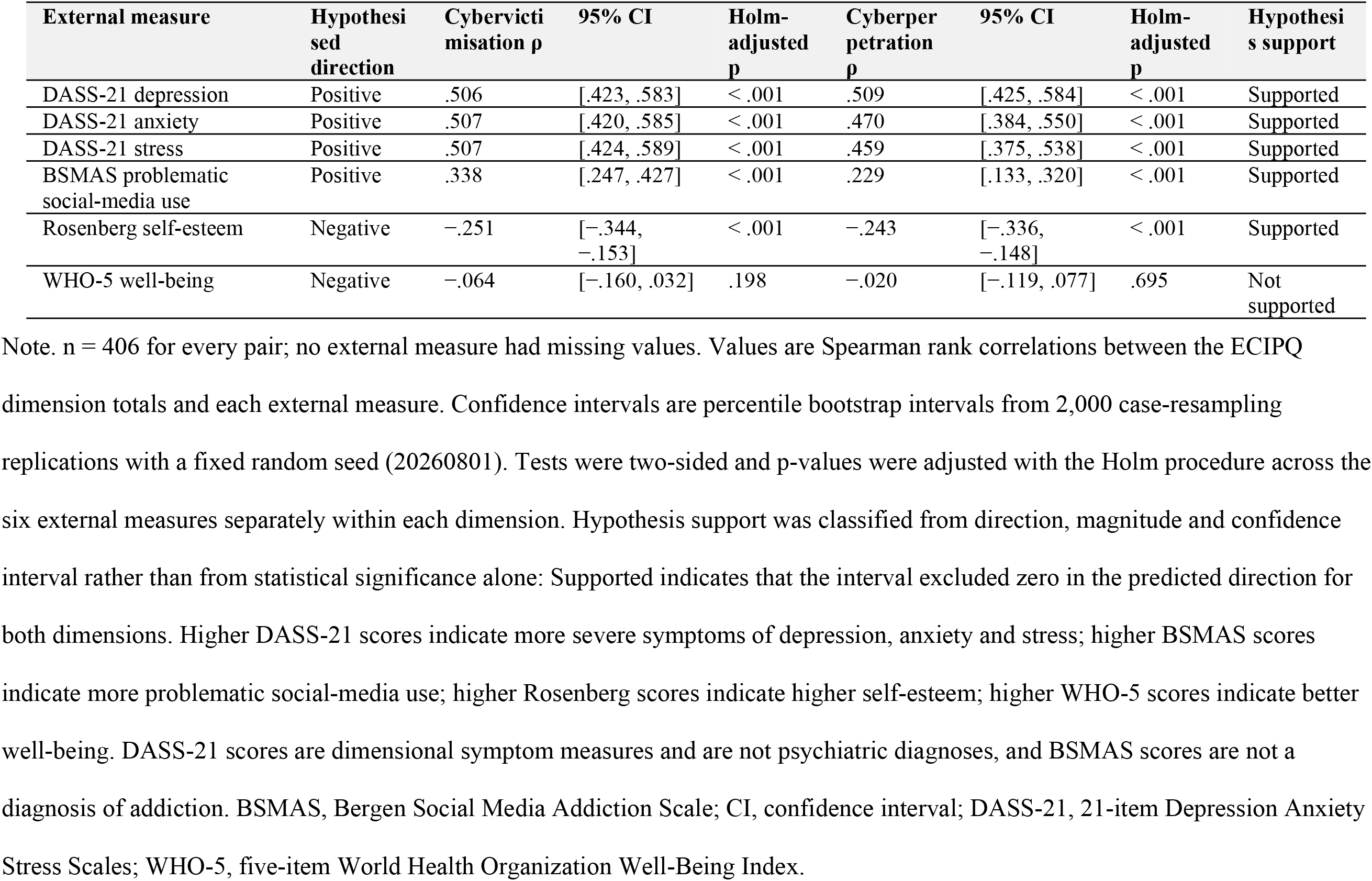
Associations of the ECIPQ dimensions with external validity measures.

### Measurement invariance across sex

The instrument operated equivalently in female and male students (Table 5). Single-group models were interpretable in both sexes, fitting well among women (CFI =.984, TLI =.982, RMSEA =.049, SRMR =.069; loadings.729 to.936; φ =.851) and among men (CFI =.988, TLI =.986, RMSEA =.049, SRMR =.053; loadings.538 to.939; φ =.831). The configural model fitted well (CFI =.986, TLI =.984, RMSEA =.049, 90% CI.040 to.057, SRMR =.061), supporting configural invariance. Constraining all item thresholds to equality across sex did not worsen fit (scaled Δχ²(22) = 20.11, p =.576; ΔCFI =.000, ΔRMSEA = −.001), so threshold invariance was supported. Additionally constraining all 22 loadings again left fit essentially unchanged (scaled Δχ²(20) = 19.29, p =.503; ΔCFI =.001, ΔRMSEA = −.002), so loading invariance was supported. No parameter required release and no partial-invariance model was needed (S9 Table). Sex-specific parameters are given in S8 Table.

**Table 5.**
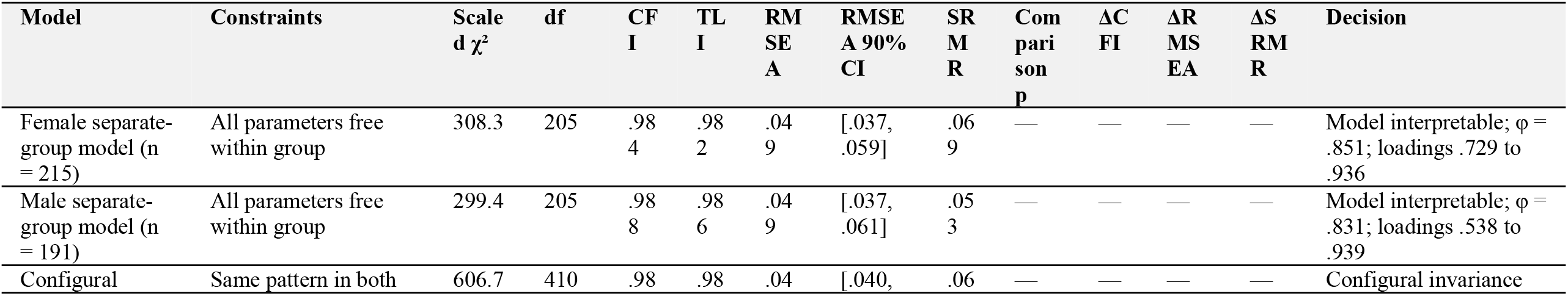
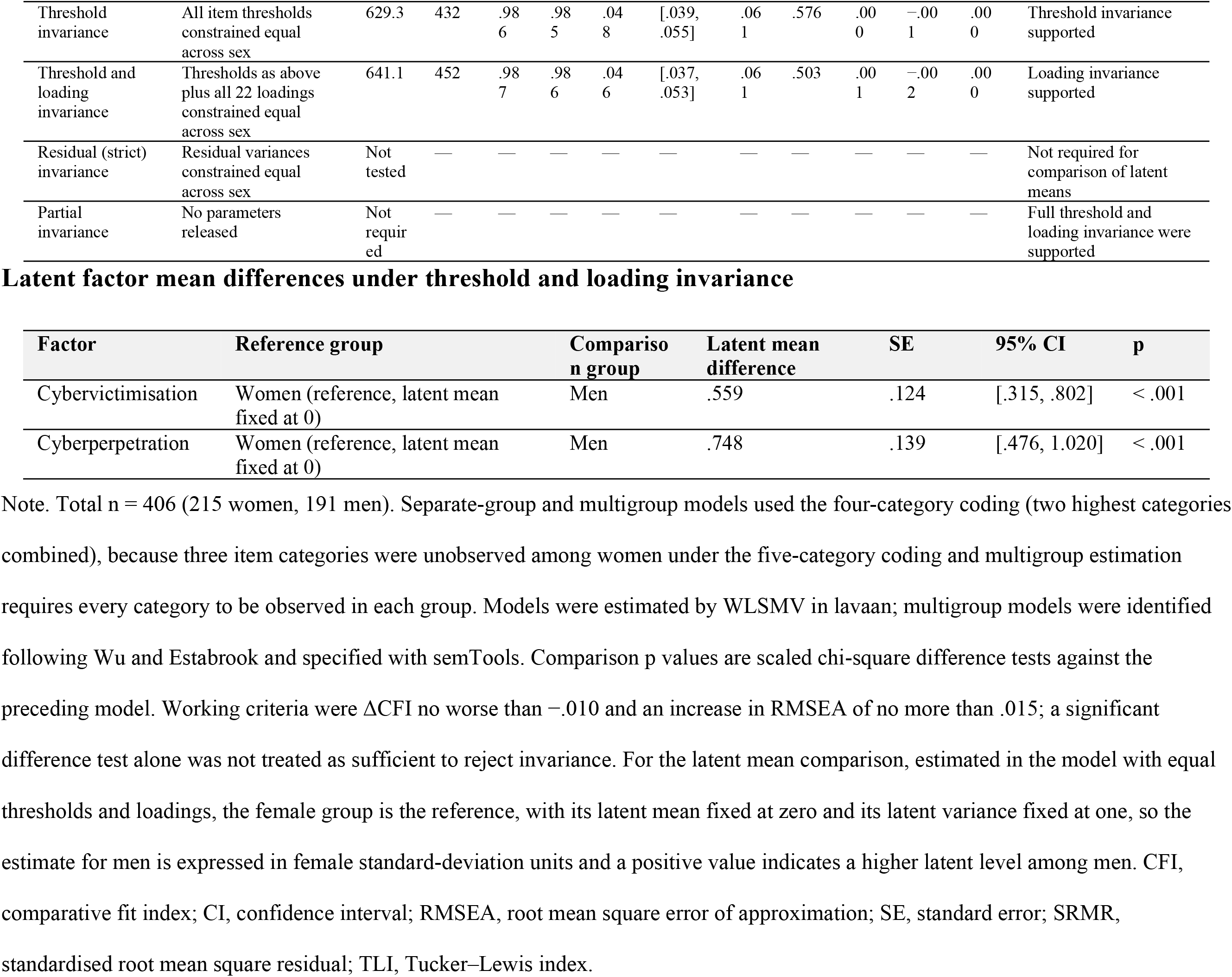
Measurement invariance of the ECIPQ-based measure across female and male students.

Because threshold and loading invariance were established, latent means could be compared. Men showed higher latent levels than women on both dimensions: the latent mean difference was.559 female standard-deviation units for cybervictimisation (95% CI.315 to.802, p <.001) and.748 units for cyberperpetration (95% CI.476 to 1.020, p <.001). These estimates are reported here as evidence that the instrument supports such comparisons; their substantive interpretation lies outside the scope of a measurement paper.

### Sensitivity analyses

The psychometric conclusions were robust across the sensitivity analyses undertaken (S10 Table). Collapsing the two highest response categories reproduced the five-category solution closely (CFI.984 versus.989; RMSEA.049 versus.045; factor correlation.849 versus.848). Excluding the single participant with an identical non-zero response to all 22 items changed no conclusion (CFI.984 versus.987; factor correlation.849 versus.839). The three reliability estimators agreed to within.003. Two analyses altered the picture in informative ways. Treating the items as continuous with robust maximum likelihood attenuated the loadings (range.508 to.845 versus.627 to.928), lowered the factor correlation (.807 versus.849) and fitted poorly (CFI =.872, RMSEA =.097), confirming that ordinal estimation is necessary for items distributed as these are. Excluding the 128 participants who scored zero on every item (four-category coding, because several upper categories became empty) weakened fit slightly and lowered the factor correlation and the smallest loading (CFI.981; RMSEA.052; factor correlation.785 versus.849; lowest loading.416 versus.627); these are valid uninvolved respondents whose exclusion restricts the range of the latent variables, so this specification is reported for completeness only and was not adopted. The supplementary bifactor model converged without improper estimates but fitted less well than the accepted model (CFI =.980, RMSEA =.057), with general-factor loadings of.557 to.878 and specific-factor loadings of.020 to.479. Latent external-validity modelling was not undertaken, for the reasons given in the Methods.

In summary, the accepted model was the theory-adjusted correlated two-factor ordinal model with three content-matched residual covariances; all 22 items were retained; configural, threshold and loading invariance across sex were supported, with residual invariance not identified in this parameterisation; and, because the correspondence between the administered items and the published ECIPQ could not be verified from the available records, the measure is described throughout as an ECIPQ-based instrument rather than as the ECIPQ itself.

## Discussion

In 406 health-professions students at Kenya Medical Training College, a 22-item ECIPQ-based measure of cybervictimisation and cyberperpetration performed well as a two-dimensional ordered categorical instrument. A correlated two-factor model fitted markedly better than a single general factor, and after three content-matched residual covariances were released the model fitted well on conventional indices. All 22 items loaded substantially on their intended factor, both dimensions were highly reliable, and configural, threshold and loading invariance across female and male students were supported. Convergent validity was supported against distress, problematic social-media use and self-esteem but not against well-being, and discriminant validity was only partially supported because the two dimensions shared most of their variance. No item warranted deletion, so the complete item set is retained.

### Structural validity

The two-dimensional result aligns with the original European validation and with subsequent work in Colombian, Chinese and Spanish samples [8–11]. That the same broad structure emerges in Kenyan health-professions students — emerging adults rather than school-aged adolescents, in a sub-Saharan African rather than a European, Latin American or East Asian setting — is a meaningful extension of the instrument’s evidence base. Two differences from earlier work deserve comment. First, the latent correlation of.849 observed here is at the upper end of published estimates, and parallel analysis retained only a single factor. Second, the loadings were uniformly high, and the reliability coefficients exceeded.95 for both dimensions, substantially above the omega values of.82 for victimisation and.68 for aggression reported in the largest recent Spanish adolescent study [11]. Several features of the present study plausibly contribute: the use of polychoric correlations and a categorical estimator rather than continuous methods, which recovers larger loadings for skewed items [13,14,43]; an older and more homogeneous sample; and the retention of the complete 22-item set. We cannot determine from these data which of these explanations predominates, and none should be treated as established.

### Factor correlation and local dependence

The strong latent correlation and the single-factor parallel-analysis result point the same way: a general involvement dimension accounts for much of the shared variance among these items in this population. This is conceptually intelligible. The ECIPQ items are constructed as matched pairs describing the same behaviour from the target and actor perspectives, so students who are involved in cyberbullying at all tend to endorse items from both sets, and the bully–victim overlap documented across the cyberbullying literature is substantial [3]. The supplementary bifactor solution was consistent with this reading, fitting less well than the correlated two-factor model and producing small specific loadings for several items (7 of 22 below.30), a pattern that typically indicates limited unique dimension-specific variance [61]. The three released residual covariances reinforce the point rather than undermining it: two of the three link matched victimisation–perpetration pairs describing exactly the same behaviour (public ridicule; insult and name-calling), and the third links two victimisation items whose content overlaps. These modifications were made after estimation, were limited to three, and were justified by documented content rather than by fit improvement alone; nonetheless they remain post hoc and require confirmation in an independent sample. The practical implication is that the two subscales should be reported separately, as the model comparison supports, while researchers should expect them to behave similarly and should not treat them as independent constructs.

### Item distributions

Every item showed a pronounced floor effect, with between 63.5% and 84.7% of students selecting never and all medians at zero. Under conventional criteria such distributions would raise concern about item quality [36], but that reading would be mistaken here. These items ask about harmful behaviours that are genuinely uncommon in a general student population, so a low base rate is the expected finding rather than a defect, and the corrected item–total correlations of.512 to.803 confirm that the items continued to discriminate. What the distributions do require is appropriate estimation: with medians of zero and sparse upper categories, mean-based summaries would be misleading if used inferentially, and treating the items as continuous attenuated the loadings in our own sensitivity analysis. The distributions also constrain the invariance analysis, since the highest thresholds are estimated with limited precision and three item categories were unobserved among women. This is why the four-category coding was used for the multigroup models, and the close agreement between the five-category and four-category single-group solutions supports that choice.

### Reliability and item retention

Both dimensions showed internal consistency well above conventional thresholds, and the three estimators agreed closely. We report omega as the principal coefficient because alpha assumes essential tau-equivalence and is a poor summary when a model-based estimate is available [51,52], and we deliberately avoid a single 22-item coefficient, which would misrepresent a two-dimensional instrument. High internal consistency should not be over-read. Coefficients of this magnitude partly reflect the general involvement dimension and the concentration of responses at the floor, and they raise the possibility of content redundancy among matched items. They are not evidence of unidimensionality, of structural validity or of construct validity, all of which rest here on the model comparison, the loading pattern, the invariance analysis and the external correlations. Because no loading fell below.40, no deletion improved dimension reliability and no item behaved inconsistently across sex, the complete content domain was preserved. Retaining all 22 items also keeps the instrument comparable with the international literature, which matters more for a measure whose principal value is cross-study comparability than a marginal gain in parsimony would.

### Convergent validity

Both dimensions correlated moderately and positively with depression, anxiety and stress symptoms and with problematic social-media use, and negatively with self-esteem, in line with the associations reported for cyberbullying involvement across the international literature [3,4] and in nursing and health-professions student samples [17]. These are cross-sectional associations and no causal interpretation is warranted in either direction. The negative association with self-esteem, while clearly present, was modest in magnitude. The clearest departure from expectation was well-being: correlations with the WHO-5 were essentially zero for both dimensions and remained non-significant after adjustment. Several readings are possible. The WHO-5 measures positive subjective well-being over the preceding two weeks rather than symptom burden, and positive and negative affect are not simple opposites, so an instrument capturing the presence of good feeling may be less sensitive to a low-frequency adverse exposure than symptom scales are [35]. The two-week reference window is also shorter than the reference period for the cyberbullying items. We cannot adjudicate between these explanations, and the finding is reported as a limitation of convergent validity rather than explained away.

### Discriminant validity and sex invariance

Integrating five sources of evidence, discriminant validity is best classified as partially supported. The model comparison, the confidence interval for the latent correlation excluding unity, the heterotrait–monotrait ratio below.90 and the observed-score correlation of.655 all support two distinguishable dimensions; the failure of the Fornell–Larcker criterion for victimisation and the single-factor parallel-analysis result qualify that conclusion. Reliance on any one criterion would have produced a different and less defensible verdict [55,56]. The invariance findings are more straightforward. Configural, threshold and loading invariance were supported and required no partial-invariance model. This is a stronger result than the configural-only invariance reported in the largest recent Spanish adolescent validation [11], and it means that scores can be compared between female and male students in this population without item-level bias. The latent mean differences observed here are consequently interpretable as differences in the underlying constructs rather than as measurement artefacts. Residual invariance was not tested and is not a prerequisite for comparing latent means.

### Implications

For research in Kenyan higher education, these findings support using the instrument with its complete 22-item set and scoring it as two separate subscales rather than as a single total, while reporting the latent correlation so that readers can judge how far the dimensions diverge in a given sample. Because measurement is invariant across sex, studies in comparable populations can compare women and men directly without first re-establishing invariance, although researchers examining other groupings — campus, faculty, programme or year of study — should not assume that invariance extends to them. For multicampus surveys, the close agreement between the five-category and four-category codings offers a practical fallback where sparse upper categories threaten estimation. For health-professions education, the associations with distress and with problematic social-media use suggest that cyberbullying is worth measuring within existing student support and counselling structures, though the instrument is a research measure of behavioural frequency and is not a diagnostic or clinical screening tool, and should not be used as one. Methodologically, the attenuation observed when items were treated as continuous is a concrete argument for categorical estimation whenever cyberbullying items with low base rates are modelled.

### Strengths and limitations

The study has several strengths. The sample was drawn from multiple campuses distributed across the country, the sex groups were close to balanced, and data were complete for all 22 items and all external measures in all 406 participants. The complete item set was modelled as ordered categorical indicators using polychoric correlations and a robust categorical estimator, the model comparison was prespecified, three reliability coefficients and five forms of discriminant-validity evidence were reported rather than a single favourable statistic, measurement invariance was tested with an identification strategy appropriate to ordered categories, and every model modification, substitution and non-undertaken analysis is documented explicitly.

The limitations are equally clear. The design is cross-sectional, so no causal inference is possible from the validity correlations, and no test–retest or longitudinal invariance evidence is available. All measures were self-reported and collected in a single sitting, so recall bias, social-desirability bias and common-method variance may all have influenced the associations reported, most plausibly by inflating them. The pronounced floor effects and sparse upper categories mean that the highest thresholds are imprecisely estimated, and three item categories were unobserved among women under the five-category coding; the invariance analysis therefore relied on the four-category coding, which mitigates but does not remove this constraint. The subsample sizes for the multigroup models (215 women, 191 men) are modest for a 22-indicator two-factor model, and while the models converged without improper estimates, the invariance conclusion would benefit from replication in a larger sample [28]. Invariance across campus, faculty and year of study was not examined. No behavioural or informant-based criterion was available, so criterion and predictive validity remain untested. The three residual covariances were released post hoc. Documentation of the instrument’s provenance and of any adaptation procedure was not available, so semantic equivalence with the published ECIPQ could not be assessed and no cognitive interviewing was conducted to establish how Kenyan students interpret the items [62]. Finally, participants were students at participating KMTC campuses and the findings should not be read as representative of Kenyan students nationally, of students at other institutions, or of non-student young adults.

### Future research

Independent replication in other Kenyan institutions, including universities and non-health programmes, would establish how far these findings generalise. Priorities include test–retest reliability and longitudinal invariance; invariance across campus, faculty and year of study in samples large enough to support it; cognitive interviewing and qualitative work on how students interpret individual items in this setting; item-response-theory analysis, which is well suited to the sparse upper categories observed here; and criterion and predictive validity against outcomes measured independently of self-report. Confirmation of the three residual covariances in an independent sample would establish whether the local dependence observed here is a stable property of the paired item design. Assembling and publishing the instrument’s provenance and adaptation record would allow a future study to state directly whether the measure is the ECIPQ or an adapted derivative of it.

## Conclusion

Among health-professions students at Kenya Medical Training College, a 22-item ECIPQ-based measure functioned as a reliable two-dimensional instrument. A correlated two-factor model representing cybervictimisation and cyberperpetration was clearly preferable to a single general factor, all 22 items loaded substantially on their intended dimension and were retained, and both dimension scores showed high internal consistency. Construct validity was supported against measures of psychological distress, problematic social-media use and self-esteem, but not against well-being. Discriminant validity was partially supported: the two dimensions were reliably distinguishable but strongly correlated, and a general involvement factor accounted for much of their shared variance.

Measurement was invariant across female and male students at the configural, threshold and loading levels, so scores can be compared between women and men in this population without item-level bias. The evidence supports using the complete 22-item set and reporting the two dimensions as separate but related subscales in research applications; it does not support use as a diagnostic or clinical screening instrument. Because the correspondence between the administered items and the published ECIPQ could not be verified from available records, the measure should be described as ECIPQ-based until provenance documentation is available. Independent replication, test–retest and longitudinal evidence, and validation against criteria measured independently of self-report remain necessary before these conclusions can be extended to other Kenyan student populations.

## Data Availability

All data underlying the findings are available without restriction from the Zenodo repository at https://doi.org/10.5281/zenodo.24253229, which contains the de-identified participant-level dataset, the variable codebook and the complete R analysis code reproducing every reported estimate.

## Acknowledgments

The authors thank the students who took part in the study, the principals of the participating Kenya Medical Training College campuses for permission to conduct the survey, and the student support services staff who served as research assistants during data collection.

## Supporting information

**S1 Checklist. STROBE checklist for cross-sectional studies.** Completed checklist indicating the section of the manuscript in which each item is addressed.

**S1 Table. Dataset and analysis-input verification.** Prespecified expectations, observed results and pass status for every data-integrity check performed before modelling.

**S2 Table. Kaiser–Meyer–Olkin and Bartlett factorability diagnostics.** Overall and item-level measures of sampling adequacy and the test of sphericity, both computed on the polychoric correlation matrix.

**S3 Table. Polychoric parallel-analysis results. Observed eigenvalues against the mean and 95th-percentile eigenvalues from 1,000 permuted datasets.**

**S4 Table. Complete item thresholds and CFA parameter estimates for the accepted model. Standardised loadings with robust standard errors, z values, residual variances and all four thresholds per item, together with the factor correlation and the three residual correlations.**

**S5 Table. Item-deletion and reliability diagnostics. Corrected item–total correlations and ordinal alpha with each item removed, with the retention decision for every item.**

**S6 Table. Complete ECIPQ item wording and analytical labels.** Full administered wording, dimension, hypothesised factor, response range and the short label used in tables and figures for each of the 22 items.

**S7 Table. Discriminant-validity evidence.** The five converging approaches used to evaluate the separability of the two dimensions, with estimates, comparators and an integrated interpretation.

**S8 Table. Sex-specific item loadings and factor correlations. Standardised loadings and factor correlations from the accepted model estimated separately in women and men (four-category coding), with the multigroup fit statistics.**

**S9 Table. Parameters released in partial-invariance models.** Records that no parameter was released because full threshold and loading invariance were supported.

**S10 Table. Sensitivity-analysis summary.** Every prespecified, conditional and exploratory sensitivity analysis, including those not undertaken and the reason.

**S11 Table. Modification indices and local-dependence diagnostics. The 15 largest residual-covariance modification indices from the prespecified two-factor model, the largest absolute residual correlations before and after modification, and the content justification for the three released parameters.**

**S12 Table. Polychoric correlation matrix of the 22 ECIPQ items. Full 22 by 22 matrix estimated by the two-step procedure with thresholds from the marginal distributions.**

**S13 Table. Item content, matched behaviours and instrument correspondence.** Within-study content mapping of the 11 matched victimisation–perpetration pairs and the assessment of correspondence with the published instrument.

**S14 Table. Reproducibility record. Software and package versions, estimator specification, parameterisation, identification, random seeds and every analytical decision.**

**S1 Fig. Parallel analysis of the 22 ECIPQ items. Observed polychoric eigenvalues plotted against the mean and 95th-percentile eigenvalues from 1,000 permuted datasets that preserve each item’s marginal distribution.**

**S2 Fig. Comparative fit of principal and sensitivity models.** Panel A plots CFI and TLI (higher is better) and Panel B plots RMSEA and SRMR (lower is better) for the models reported in Table 2, so that indices with opposite desirable directions are not combined on a single scale. The bifactor and continuous-item models are excluded because their unadjusted statistics are not comparable.

**S1 File. Complete R analysis code reproducing every result reported in the manuscript and supporting information.**

